# Association between individual and country-level socioeconomic factors and work participation in spondyloarthritis including psoriatic arthritis: analysis of the ASAS-perSpA study

**DOI:** 10.1101/2021.05.29.21257923

**Authors:** Sizheng Steven Zhao, Elena Nikiphorou, Annelies Boonen, Clementina López-Medina, Maxime Dougados, Sofia Ramiro

**Affiliations:** Musculoskeletal biology, Institute of Life Course and Medical Sciences, University of Liverpool, Liverpool, UK; Centre for Rheumatic Diseases, King’s College London, London, UK; Department of Rheumatology, King’s College Hospital, London, UK; Maastricht UMC+, Dept. of Internal Medicine, Div. of Rheumatology, Maastricht, the Netherlands; Care and Public Health Research Institute (CAPHRI), Maastricht University, Maastricht, the Netherlands; Université de Paris, Rheumatology Department, Cochin Hospital, Paris, France; Rheumatology Department, Reina Sofia Hospital, IMIBIC, University of Cordoba, Cordoba, Spain; INSERM (U1153): Clinical Epidemiology and Biostatistics, University of Paris, Paris, France; Department of Rheumatology, Leiden University Medical Center, Leiden, The Netherlands; Department of Rheumatology, Zuyderland Medical Center, Heerlen, the Netherlands

**Author notes:** Correspondence to: Sofia Ramiro, Department of Rheumatology, Leiden University Medical Center, Albinusdreef 2, 2333 GA Leiden, the Netherlands, P.O. Box 9600, 2300RC Leiden, the Netherlands.

**Keywords:** spondyloarthritis, psoriatic arthritis, work participation, education, healthcare expenditure, socioeconomic factors

## Abstract

**Objective:** To examine whether associations between socioeconomic factors and work outcomes in spondyloarthritis (SpA) differ across axial (axSpA), peripheral SpA (pSpA) and psoriatic arthritis (PsA), and whether associations for individual-level socioeconomic factors are modified by country-level factors.

**Methods:** Patients with a physician diagnosis of SpA within working age (18-65 years) were included. Associations between individual- (age, gender, education, marital status) and country-level factors (Human Development Index, Health Care Expenditure (HCE), Gross Domestic Product, percentage unemployed) with work outcomes (employment status, absenteeism, presenteeism) were assessed using multivariable mixed-effects models. Associations between individual factors and outcomes were compared according to SpA phenotypes and country-level factors using interaction terms.

**Results:** A total of 3835 patients (mean age 42 years, 61% males) from 23 countries worldwide were included (66% axSpA, 10% pSpA, 23% PsA). Being employed was associated with gender (male vs. female OR 2.5; 95%CI 1.9-3.2), education (university vs. primary OR 3.7; 2.9-4.7), marital status (married vs. single OR 1.3; 1.04-1.6), and age in a non-linear manner. University (vs primary) education was associated with lower odds of absenteeism (OR 0.7; 0.5-0.96) and presenteeism (OR 0.5; 0.3-0.7). Associations were similar across SpA phenotypes. Higher HCE was associated with more favourable work outcomes, e.g., higher odds of employment (OR 2.5; 1.5-4.1). Gender discrepancy in odds of employment was greater in countries with lower socioeconomic development.

**Conclusion:** Higher educational attainment and higher HCE were associated with more favourable work outcomes, independently of SpA phenotype. The disadvantageous effect of female gender on employment is particularly strong in countries with lower socioeconomic development.

## Introduction

Spondyloarthritis (SpA) comprises a heterogenous group of disorders that share common clinical, genetic, and pathophysiological features [1]. SpA with predominantly peripheral involvement is classified as peripheral SpA (pSpA) and includes psoriatic arthritis (PsA), while axial SpA (axSpA) has a predilection for the spine and sacroiliac joints. Despite being closely related, research into these members of the SpA family have developed at different paces. One important example is work participation. Evidence for the impact of SpA on work participation and associated productivity costs comes mainly from axSpA [2–4]. Despite high levels of work disability reported in PsA, most studies were small and very few have examined associated factors [5]. Furthermore, how work participation in PsA relates to the wider context of axSpA or pSpA is unclear.

In axSpA, work disability is associated with clinical factors such as high disease activity and functional impairment – factors that can be modified (at least in part) by treatment thus representing a potential target for supporting patients to remain productive at work [6]. However, variability in treatment access and wider health inequalities exist worldwide [7]. There are remarkable differences in work status and productivity costs even between neighbouring European countries despite comparable quality of care for chronic diseases [3,4]. Worldwide, country-level socioeconomic factors (e.g., healthcare expenditure) are associated with employment in SpA [8]; these often overlooked contextual factors are essential for understanding widespread inequality in disease outcomes. A prior study reported associations between individual-level factors (e.g., gender and education) and employment status [8], but how country-level factors modify the effect of individual-level socioeconomic factors remains an unmet research need.

The main aim of this study was to understand the association between individual- and country-level socioeconomic factors and work outcomes in SpA including PsA. In particular, we examined whether these associations differed by disease phenotype (axSpA, pSpA and PsA), and whether associations observed for individual-level socioeconomic factors were modified by country-level factors.

## Methods

### Patients and study design

The ASAS-perSpA (peripheral involvement in SpA) study design has been described previously [9]. It was an international, cross-sectional study with 24 contributing countries that recruited from 2018-2020. Adult patients with a rheumatologist diagnosis of SpA were consecutively enrolled. The study was approved by the ethical committees in all countries and written informed consent was obtained from all subjects. This analysis was restricted to patients with a physician diagnosis of axSpA, PsA or pSpA between ages of 18 and 65 (i.e. working age) who completed the Work Productivity and Activity Impairment questionnaire General Health (WPAI-GH [10]).

### Outcomes

Work participation was assessed using four outcomes derived from the WPAI-GH: current employment status (being employed or not); absenteeism in those employed (i.e., percent time absent from work, derived from hours missed due to health problems, hours missed due to other reasons, and hours actually worked); presenteeism (percentage derived from degree health affected productivity while working); and percent overall work impairment due to health (absenteeism plus proportion of hours worked accounting for any impaired productivity).

The three percentage outcomes were categorised to accommodate their skewed distribution: absenteeism as 0%, 1-20%, >20% (i.e., 0 plus two equal quantiles); presenteeism as 0-10%, 20-30%, ≥40% (tertiles); overall work impairment as 0-10%, 11-40%, >40% (tertiles). Only participants in employment were included for analysis of absenteeism, presenteeism and overall work impairment. Participants reporting 100% absenteeism were excluded from analyses of presenteeism.

### Individual- and country-level socioeconomic factors

Individual socioeconomic factors comprised age, gender, education (primary, secondary, university), and marital status (single, married, divorced/widowed).

The four country-level socioeconomic factors were: Human Development Index (HDI) which ranks countries according to life expectancy, education and gross national income; Health Care Expenditure (HCE) as percentage of Gross Domestic product (GDP); GDP (based on purchasing power parity); economic unemployment (as percentage of total labour force). Latest values of each factor were obtained from the World Bank (i.e., 2018, except HCE which was only available for 2017) [11]; details are provided in supplementary Table S1. Each factor was dichotomised by the median to accommodate skewed distributions and facilitate interpretation of results.

### Covariates

Covariates tested in regression models included: body mass index (BMI), smoking status (ever/never), HLA-B27 (positive/negative/missing), axial involvement, peripheral involvement; history of enthesitis, dactylitis, uveitis, psoriasis, inflammatory bowel disease (IBD); Bath AS disease activity index (BASDAI), CRP, AS disease activity score (ASDAS), fatigue (BASDAI question 1); depression or anxiety (present or absent according to the three-level version of the EQ-5D question 5, henceforth abbreviated as ‘depression’), fibromyalgia (physician diagnosis); NSAIDs (in the past month), conventional synthetic (csDMARDs), biologic DMARDs (bDMARDs) (both since diagnosis), and current glucocorticoid use.

### Statistical analyses

The associations between individual socioeconomic factors and work outcomes were tested in a series of multivariable mixed-effects models, one for each work outcome. Mixed-effects models account for within-country similarities in the estimation of between-country differences, by incorporating country-specific means (i.e., random intercepts for each country). Logistic mixed-effects model was used for employment and ordinal logistic for categorised absenteeism, presenteeism, and overall work impairment. Each work outcome was the dependent variable and all four individual socioeconomic factors (age, gender, education, marital status) were entered as independent variables. Non-linear effects of age were tested using polynomials (e.g., age^2^). Covariates that were univariably associated with the work outcome of interest at p<0.2 were sequentially added to the model, and retained if they significantly contributed to explain the outcome (p<0.05) or changed the estimate of the main relationships of interest by 20%. Due to collinearity, separate models were created for 1) ASDAS and fatigue, 2) BASFI and fatigue, and 3) BASDAI and log-transformed-CRP. The effect of an individual socioeconomic factor on a work outcome (i.e., the coefficient, or regression line slope) could potentially differ by country; we tested this by comparing models with and without country-specific effects (i.e., random slopes) for each individual socioeconomic factor, using the likelihood ratio test.

We added interaction terms between each individual socioeconomic factor and disease phenotype (axSpA, PsA, pSpA) to test differential effects according to disease. Interaction terms with p<0.05 were taken to indicate significant effect modification by disease (conservative p-threshold chosen to account for multiple testing).

Each country level socioeconomic factor was added in separate models (due to collinearity) into the final model for each work outcome. Where random slopes indicated differential effects of an individual socioeconomic factor across countries, we further examined this by including an interaction term between the individual and each country-level socioeconomic factor. That is, we tested whether the association between individual socioeconomic factors and work outcomes differed according to country-level socioeconomic factors. Significant interactions were shown using models stratified by high and low levels of each country-level socioeconomic factor to facilitate interpretation. All analyses were performed using Stata v13.

## Results

A total of 3835 participants from 23 countries were eligible for this analysis (no participants from the UK due to administrative limitations [9]). The mean age was 42 years (SD 12) with 61% males. Forty-two percent reported university, 43% secondary and 15% primary school education. Twenty-nine percent were single, 64% married and 7% divorced or widowed. Country-specific characteristics are shown in supplementary Tables S2 and S3. Overall, 66% of participants had axSpA, 10% had pSpA and 23% had PsA (country-specific descriptions shown in supplementary Figure S1). AxSpA participants were younger, more frequently males, and had lower BMI than pSpA and PsA (**Table 1**).

**Table 1.**
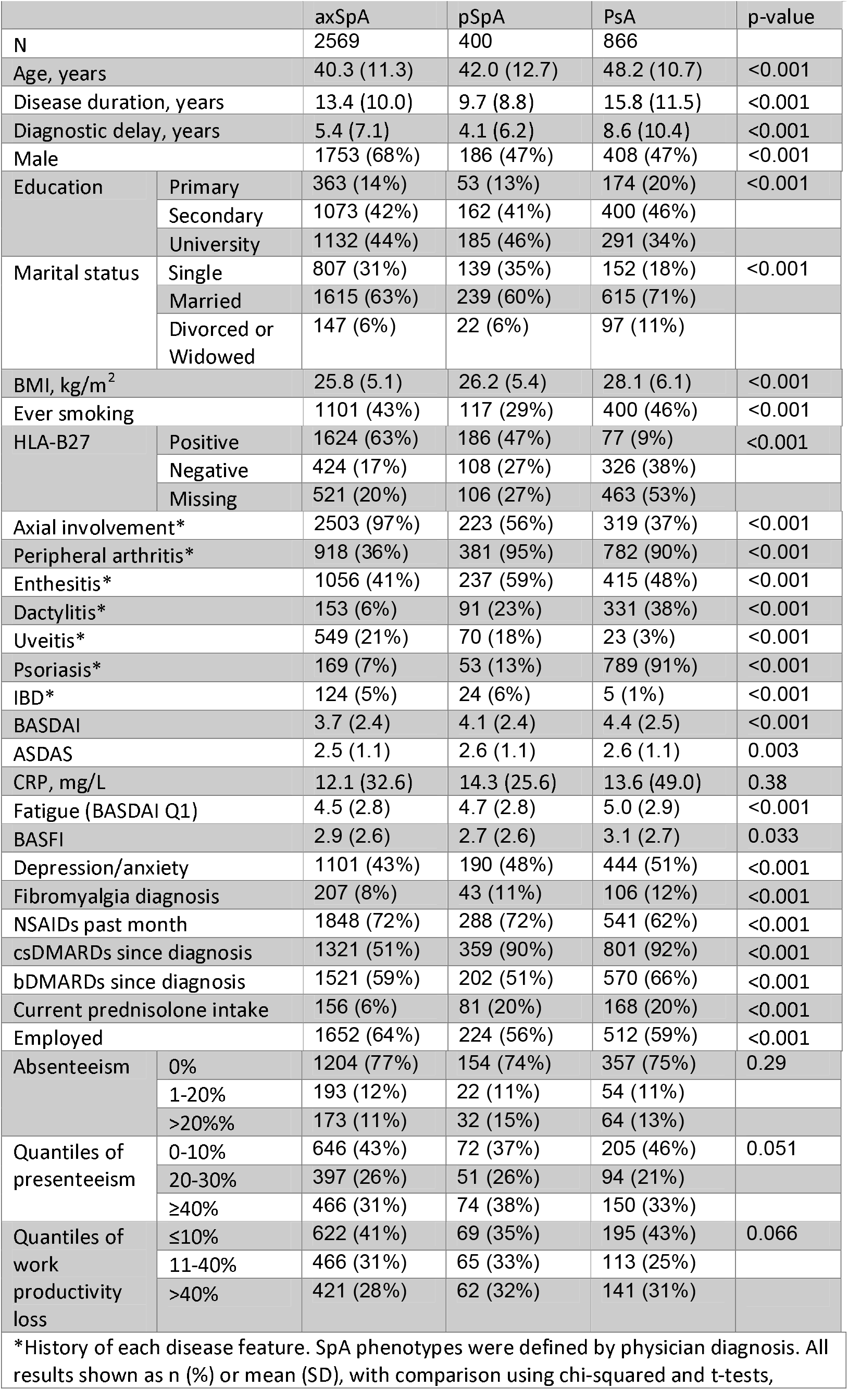
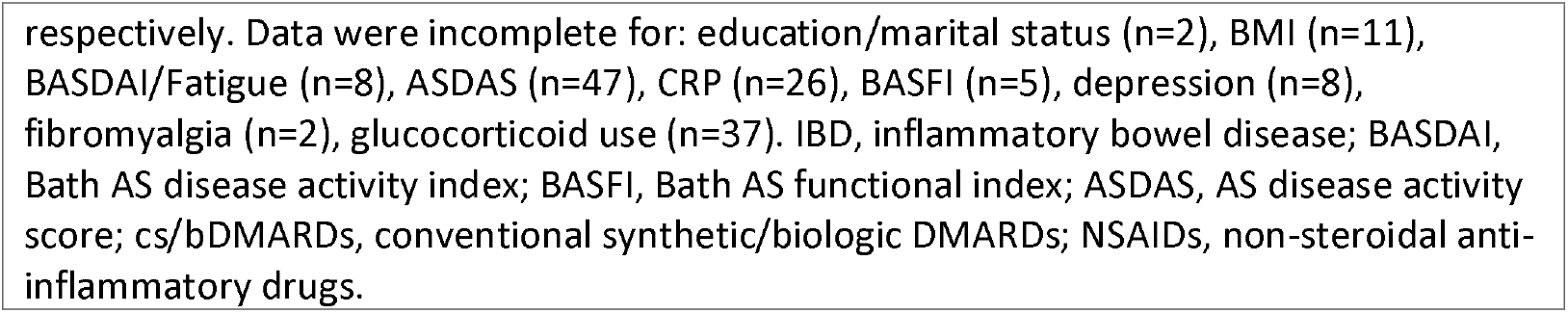
Patient characteristics according to spondyloarthritis phenotype.

|  |  |  |  |  |  |
| --- | --- | --- | --- | --- | --- |
|  |  | axSpA | pSpA | PsA | p-value |
| N |  | 2569 | 400 | 866 |  |
| Age, years |  | 40.3 (11.3) | 42.0 (12.7) | 48.2 (10.7) | <0.001 |
| Disease duration, years |  | 13.4 (10.0) | 9.7 (8.8) | 15.8 (11.5) | <0.001 |
| Diagnostic delay, years |  | 5.4 (7.1) | 4.1 (6.2) | 8.6 (10.4) | <0.001 |
| Male |  | 1753 (68%) | 186 (47%) | 408 (47%) | <0.001 |
| Education | Primary | 363 (14%) | 53 (13%) | 174 (20%) | <0.001 |
|  | Secondary | 1073 (42%) | 162 (41%) | 400 (46%) |  |
|  | University | 1132 (44%) | 185 (46%) | 291 (34%) |  |
| Marital status | Single | 807 (31%) | 139 (35%) | 152 (18%) | <0.001 |
|  | Married | 1615 (63%) | 239 (60%) | 615 (71%) |  |
|  | Divorced or Widowed | 147 (6%) | 22 (6%) | 97 (11%) |  |
| BMI, kg/m<sup>2</sup> |  | 25.8 (5.1) | 26.2 (5.4) | 28.1 (6.1) | <0.001 |
| Ever smoking |  | 1101 (43%) | 117 (29%) | 400 (46%) | <0.001 |
| HLA-B27 | Positive | 1624 (63%) | 186 (47%) | 77 (9%) | <0.001 |
|  | Negative | 424 (17%) | 108 (27%) | 326 (38%) |  |
|  | Missing | 521 (20%) | 106 (27%) | 463 (53%) |  |
| Axial involvement* |  | 2503 (97%) | 223 (56%) | 319 (37%) | <0.001 |
| Peripheral arthritis* |  | 918 (36%) | 381 (95%) | 782 (90%) | <0.001 |
| Enthesitis* |  | 1056 (41%) | 237 (59%) | 415 (48%) | <0.001 |
| Dactylitis* |  | 153 (6%) | 91 (23%) | 331 (38%) | <0.001 |
| Uveitis* |  | 549 (21%) | 70 (18%) | 23 (3%) | <0.001 |
| Psoriasis* |  | 169 (7%) | 53 (13%) | 789 (91%) | <0.001 |
| IBD* |  | 124 (5%) | 24 (6%) | 5 (1%) | <0.001 |
| BASDAI |  | 3.7 (2.4) | 4.1 (2.4) | 4.4 (2.5) | <0.001 |
| ASDAS |  | 2.5 (1.1) | 2.6 (1.1) | 2.6 (1.1) | 0.003 |
| CRP, mg/L |  | 12.1 (32.6) | 14.3 (25.6) | 13.6 (49.0) | 0.38 |
| Fatigue (BASDAI Q1) |  | 4.5 (2.8) | 4.7 (2.8) | 5.0 (2.9) | <0.001 |
| BASFI |  | 2.9 (2.6) | 2.7 (2.6) | 3.1 (2.7) | 0.033 |
| Depression/anxiety |  | 1101 (43%) | 190 (48%) | 444 (51%) | <0.001 |
| Fibromyalgia diagnosis |  | 207 (8%) | 43 (11%) | 106 (12%) | <0.001 |
| NSAIDs past month |  | 1848 (72%) | 288 (72%) | 541 (62%) | <0.001 |
| csDMARDs since diagnosis |  | 1321 (51%) | 359 (90%) | 801 (92%) | <0.001 |
| bDMARDs since diagnosis |  | 1521 (59%) | 202 (51%) | 570 (66%) | <0.001 |
| Current prednisolone intake |  | 156 (6%) | 81 (20%) | 168 (20%) | <0.001 |
| Employed |  | 1652 (64%) | 224 (56%) | 512 (59%) | <0.001 |
| Absenteeism | 0% | 1204 (77%) | 154 (74%) | 357 (75%) | 0.29 |
|  | 1-20% | 193 (12%) | 22 (11%) | 54 (11%) |  |
|  | >20% | 173 (11%) | 32 (15%) | 64 (13%) |  |
| Quantiles of presenteeism | 0-10% | 646 (43%) | 72 (37%) | 205 (46%) | 0.051 |
|  | 20-30% | 397 (26%) | 51 (26%) | 94 (21%) |  |
|  | ≥40% | 466 (31%) | 74 (38%) | 150 (33%) |  |
| Quantiles of work productivity loss | ≤10% | 622 (41%) | 69 (35%) | 195 (43%) | 0.066 |
|  | 11-40% | 466 (31%) | 65 (33%) | 113 (25%) |  |
|  | >40% | 421 (28%) | 62 (32%) | 141 (31%) |  |
| *History of each disease feature. SpA phenotypes were defined by physician diagnosis. All results shown as n (%) or mean (SD), with comparison using chi-squared and t-tests, |  |  |  |  |  |

respectively. Data were incomplete for: education/marital status (n=2), BMI (n=11), BASDAI/Fatigue (n=8), ASDAS (n=47), CRP (n=26), BASFI (n=5), depression (n=8), fibromyalgia (n=2), glucocorticoid use (n=37). IBD, inflammatory bowel disease; BASDAI, Bath AS disease activity index; BASFI, Bath AS functional index; ASDAS, AS disease activity score; cs/bDMARDs, conventional synthetic/biologic DMARDs; NSAIDs, non-steroidal anti-inflammatory drugs.

Across all participants, 2388 (62%) were employed. Among the employed, the mean (SD) and median (IQR) absenteeism was 9% (SD 23%) and 0% (IQR 0-0%), presenteeism 26% (SD 26%) and 20% (IQR 0-50%), and overall work impairment 29% (SD 28%) and 20% (IQR 0-50%); 528 (24%) reported some degree (i.e., >0%) of absenteeism, 1532 (71%) some degree (>0%) of presenteeism, and 1545 (72%) some degree (>0%) of overall work impairment. Sixty-four percent of axSpA participants were employed, compared to 56% of pSpA and 59% PsA (p=0.74 after adjusting for age, gender, educational and marital status), but other work outcomes were not significantly different (**Table 1**). Participant characteristics according to employment status are shown in supplementary Table S4; briefly, the employed were younger, more often male, and had higher education than those not employed.

Unadjusted work outcomes varied markedly across countries. For example, Taiwan had the highest percentage of employed patients (80%) and Morocco the lowest (39%) (**Figure 1**). Absenteeism ranged from 0% in Canada to 22% (SD 33%) in Egypt ; presenteeism from 17% (SD 24%) in Japan to 45% (SD 24%) in Columbia; overall work impairment from 18% (SD 24%) in Japan to 47% (SD 26%) in Columbia; the mean and median absenteeism, presenteeism, and overall work impairment for each country are shown in **Figure 1**.

**Figure 1.**
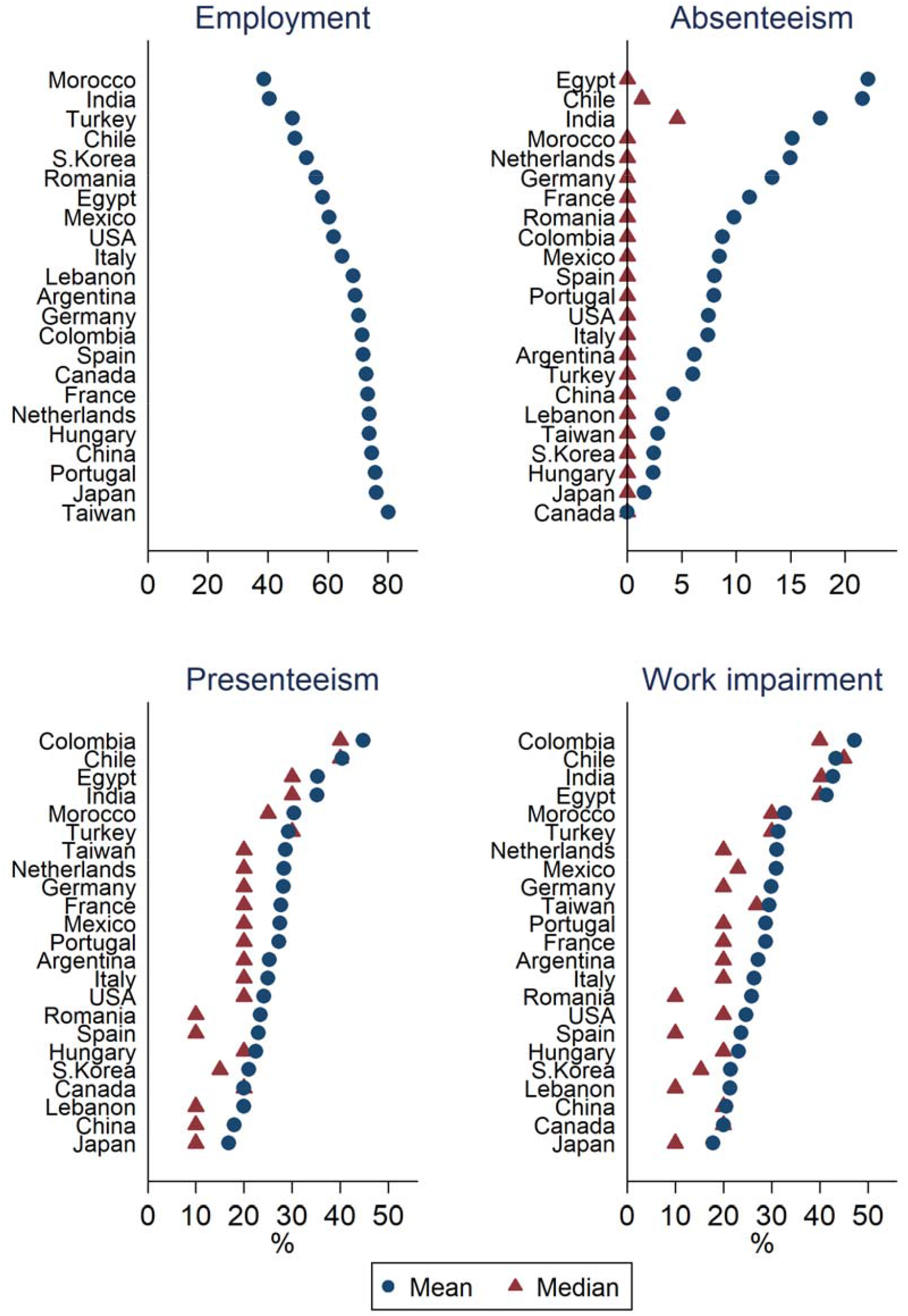
Unadjusted comparison of work outcomes between 23 countries.

### Individual-level socioeconomic factors

In multivariable mixed-effects models, odds of employment were 2.5-fold higher in males than females, 3.7-fold higher in those with university than primary education, and 27% higher in married than single participants (**Figure 2**). Odds of employment differed significantly according to age in a non-linear manner (here modelled as a quadratic curve); overall, odds were lower in the under-20s and over-60s, and highest between 30s and 50s. Country specific associations between age and employment using higher-degree polynomials (i.e., improved fit) are shown in supplementary Figure S2. Odds of employment were generally similar between countries at both ends of the working age range (18-65), with variation most marked in the 30-40s age range (supplementary Figure S2).

**Figure 2.**
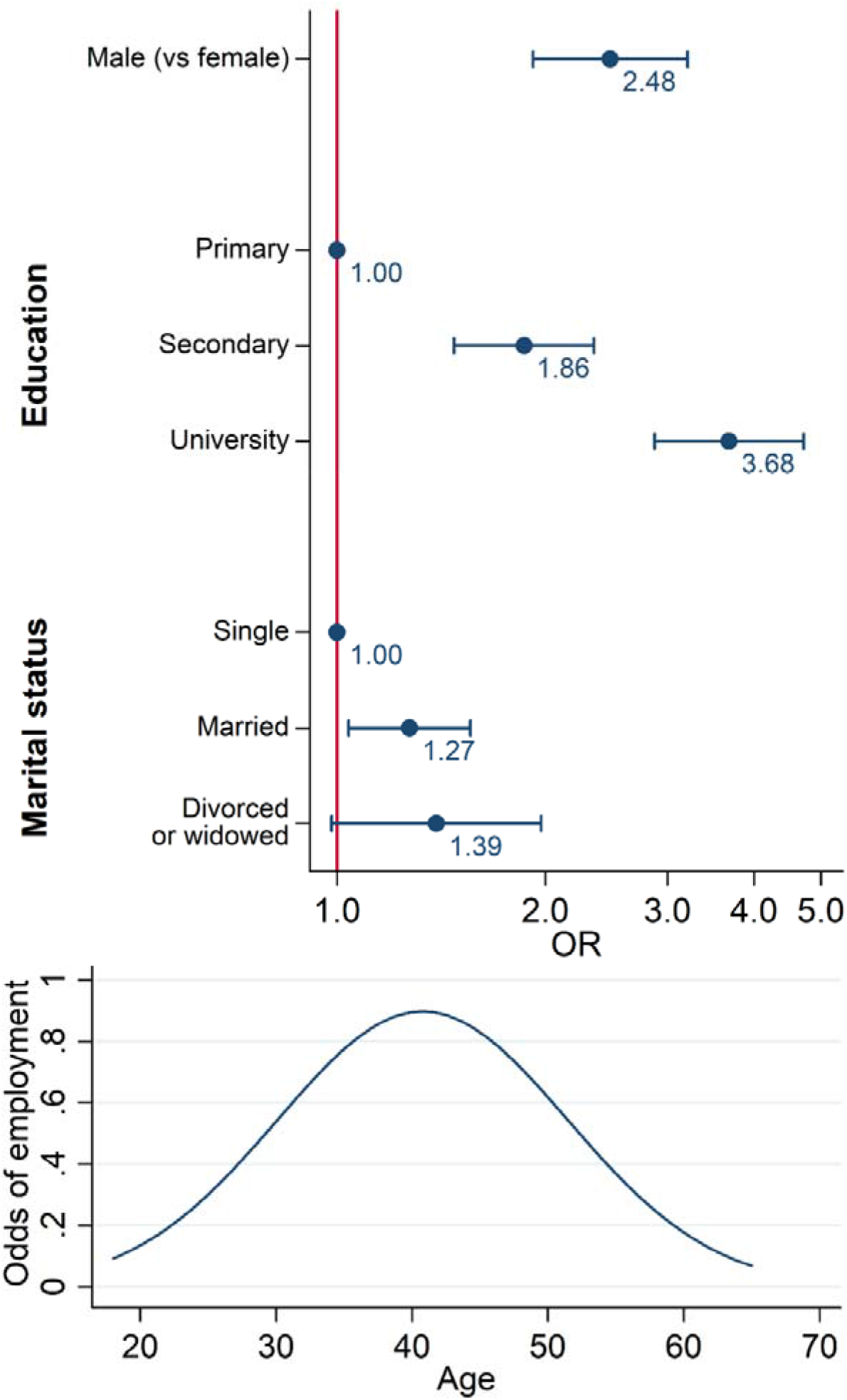
Association between individual socioeconomic factors and employment derived from multivariable mixed-effects models adjusted for ASDAS, depression and dactylitis (full model coefficients shown in supplementary Table S5). Association with age was non-linear, here modelled as a quadratic curve.

High level of education was consistently associated with lower odds of absenteeism, presenteeism and overall work impairment (**Figure 3**). Age, gender and being married had no meaningful effect on these outcomes. Full model coefficients, including for covariates, are shown in supplementary Table S5. In brief, higher ASDAS, greater fatigue, presence of depression or fibromyalgia, and NSAID use in the past month were each associated with increased absenteeism, presenteeism, and overall work impairment. Depression and ASDAS were additionally associated with reduced employment.

**Figure 3.**
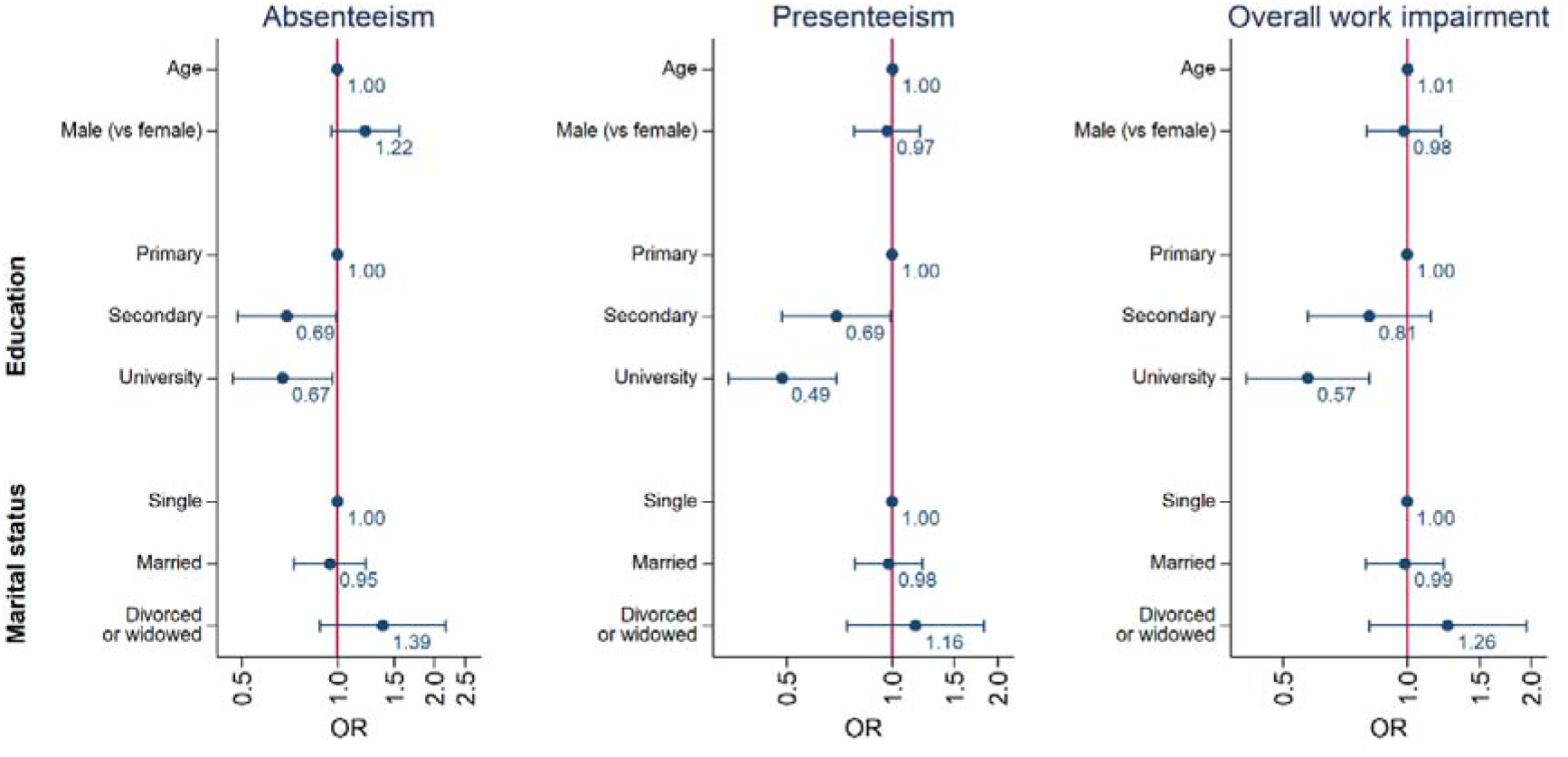
Associations between individual socioeconomic factors and absenteeism, presenteeism and overall work impairment, derived from multivariable mixed-effects models (full model coefficients shown in supplementary Table S5).

Associations between individual socioeconomic factors and work outcomes were not modified by disease phenotype; that is, interaction terms were not statistically significant (data not shown). Results did not differ significantly when using BASFI or BASDAI instead of ASDAS in the various models (supplementary Tables S6 and S7).

### Country-level socioeconomic factors

Country-level socioeconomic data for each country are described in supplementary Table S1. Higher HCE as percentage of GDP (vs lower, dichotomised by the median) was significantly associated with higher odds of employment (OR 2.5; 95%CI 1.5 to 4.1), and lower odds of absenteeism (OR 0.6), presenteeism (OR 0.6) and overall work impairment (OR 0.5) (**Table 2**). Higher HDI was similarly associated with each work outcome, except for presenteeism. The direction of effect sizes for GDP and economic unemployment were consistent with the above, but not statistically significant.

**Table 2.** Association between country-level socioeconomic factors and work outcomes.

|  |  |  |  |  |
| --- | --- | --- | --- | --- |
|  | Employment | Absenteeism | Presenteeism | Overall work impairment |
| HDI | **2.02 (1.14,3.58)** | **0.63 (0.41, 0.96)** | 0.70 (0.40, 1.21) | **0.60 (0.38,0.96)** |
| HCE (% GDP) | **2.50 (1.51,4.12)** | **0.62 (0.41, 0.94)** | **0.56 (0.33, 0.93)** | **0.54 (0.34,0.84)** |
| GDP (international \$) | 1.26 (0.67,2.36) | 0.93 (0.58, 1.49) | 0.64 (0.36, 1.12) | 0.69 (0.42,1.15) |
| Unemployment (% total labour force) | 0.80 (0.42,1.52) | 1.21 (0.76, 1.94) | **1.74 (1.03, 2.95)** | 1.60 (0.99,2.59) |
| Each country-level socioeconomic factor was added (in separate models due to collinearity) into the final multivariable mixed-effects models shown in Figures 2, 3 and Supplementary Table S4.<br>HDI, Human Development Index; HCE, Health Care Expenditure as percentage of GDP; GDP, Gross Domestic Product based on purchasing power parity in international dollars. All values from 2018, except HCE from 2017. |  |  |  |  |

### Interaction between country- and individual-level socioeconomic factors

The association between gender and employment differed across countries, that is, addition of a random slope for gender produced a statistically significant model improvement. To facilitate interpretation, we tested interaction between gender and each country-level socioeconomic factor. The gender discrepancy in employment (i.e., association between gender and employment) was greater in countries with lower levels of socioeconomic development; for example, males had 3.6-fold higher odds of employment than females in countries with low HDI, whereas odds were 1.7 in those with high HDI (**Figure 4**). Adding random slopes did not significantly improve models for the other three work outcomes (i.e., the association between the tested individual socioeconomic factors and the three work outcomes did not vary significantly across countries).

**Figure 4.**
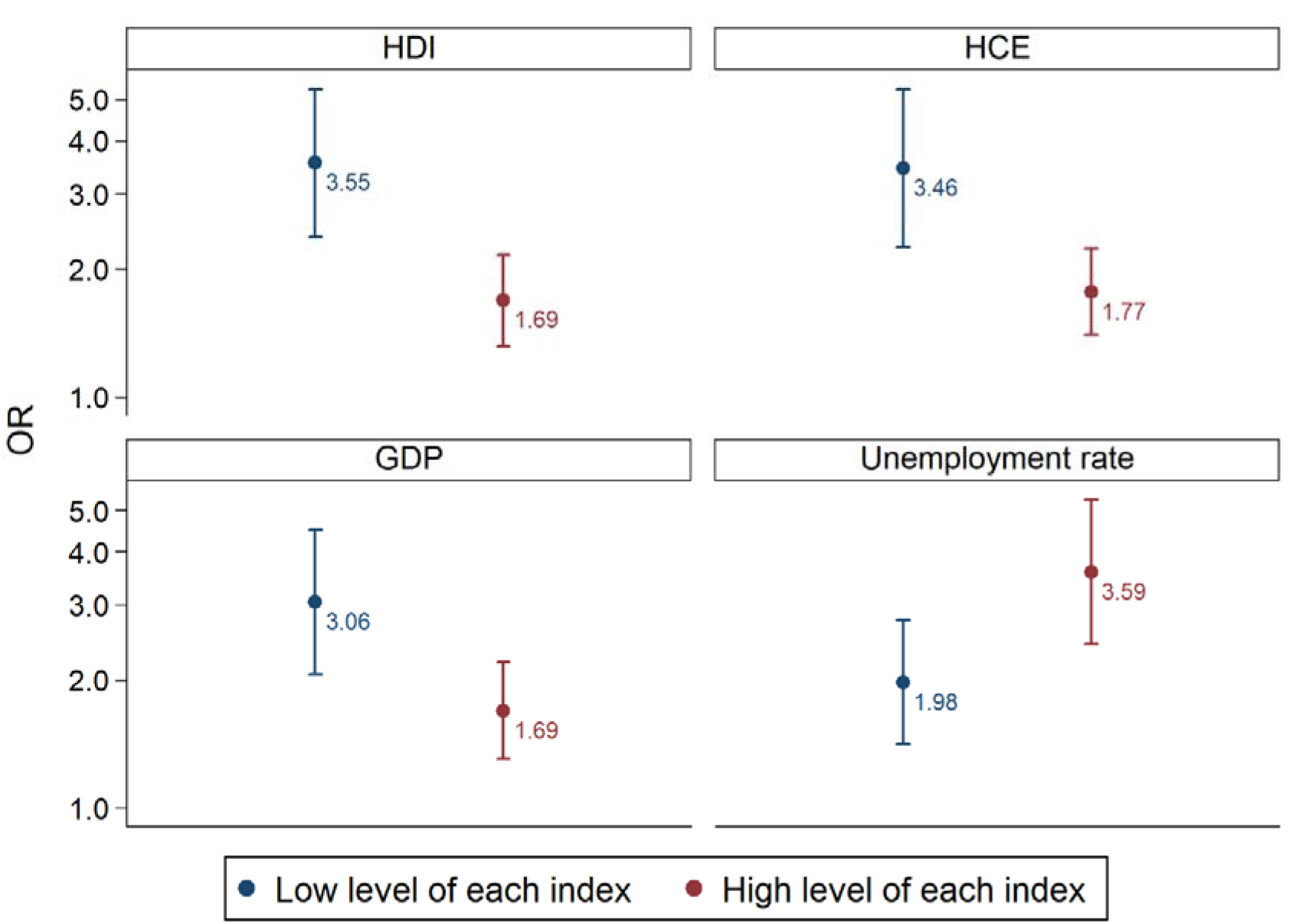
Gender differences in odds of employment differ between countries with low and high levels of socioeconomic development. Comparisons shown as males vs. females. Low and high defined by median value of each of the four country-level socioeconomic indices. For example, males had 3.55 higher odds of employment than females in countries with low HDI, whereas the difference was smaller at 1.69 fold in high HDI countries. HDI, Human development index, HCE, healthcare expenditure as proportion of GDP; GDP, gross domestic product.

## Discussion

This is one of the largest, international, cross-sectional studies in SpA and the first to compare work participation and associated factors across the SpA phenotypes. We showed that lower education and lower country-level healthcare expenditure were consistently and independently associated with poorer work participation outcomes. These associations did not differ across SpA phenotypes. The extent to which gender was associated with employment status differed between countries: the discrepancy between males (who had higher odds of employment) vs. females was greater in countries with lower (compared to higher) socioeconomic development.

As in the ASAS-COMOSPA study, we found significant between-country variation in work participation [8]. There were no significant differences in employment status, absenteeism, presenteeism, overall work impairment, or their predictive factors across SpA phenotypes. This suggests that prior research on work participation in axSpA may be extrapolatable to the rest of the SpA family. It also supports a unified approach to SpA management and research with respect to work outcomes, rather than the historically divergent efforts. Age was associated with being employed in the current study, but not in ASAS-COMOSPA; this is likely due to the need to model their non-linear relationship. We showed that odds of employment were highest in the 30-50s range; this was also the age range in which there was the greatest between-country variation in odds of employment. In the ASAS-COMOSPA study, education was only associated with employment status [8], whereas we found that lower education was also significantly associated with higher absenteeism, presenteeism, and overall work impairment among the employed, independent of disease activity. The reason for discrepant results for education is unclear, although we did additionally consider important confounders such as depression, fibromyalgia, and fatigue. Male gender was significantly associated with employment status in both studies, but we also showed that the magnitude of this association differed dramatically between countries with high and low socioeconomic development. This may partly explain conflicting reports of gender-employment association in smaller studies from different parts of the world [12]. How these gender discrepancies in employment status differ between SpA and general populations in each country can only be speculated upon. It does suggest that interventions to improve work participation should be specific to each healthcare system, but ultimately interventions at the societal level are needed. To further support this, all five measures of country-level socioeconomic development were numerically associated with each of the four work outcomes. This is the first study to demonstrate significant independent associations between country-level socioeconomic factors and absenteeism, presenteeism, and overall work impairment. Healthcare expenditure as a percentage of GDP was most consistently associated with work participation in SpA patients, suggesting that healthcare policies may be more influential than workplace policies.

Higher disease activity (whether assessed by ASDAS or BASDAI) and functional impairment (BASFI) were each associated with adverse work outcomes in respective models despite adjusting for multiple potential confounders. This highlights the need for optimal disease and symptom management, which may in turn improve work outcomes. Self-reported depression and/or anxiety was consistently associated with poorer work outcomes using all four measures of work participation, independent of disease activity. Similarly, fibromyalgia and fatigue were each significantly associated with absenteeism, presenteeism and overall work impairment. Causal interpretation should be made with caution since these factors were not primary targets of study. However, depression and fibromyalgia are highly prevalent comorbidities that have disproportional impact on the management of SpA [13–15]; future studies of work participation should focus on the potential causal contributions of these important comorbidities.

A key strength of this study is its large, multinational sample. This allowed us to examine global as well as between-country associations, thus giving the results greater generalisability to clinical practice worldwide. The robust data collection process also meant minimal missing data for most variables, thus reducing bias during analysis. There were also limitations. The cross-sectional design meant that it was difficult to unravel cause and effect; for example, depression may reduce work participation, but unemployment will in turn impact mental health. The temporality issue is less relevant for most of the individual- and all the country-level socioeconomic factors (e.g., an individual’s current employment status cannot affect historical education or HCE of the country). The proportion of participants reporting university education was high (42%), which may suggest limited representability of background populations. However, it is more likely an artefact of data collection – participants with any post-secondary education were all categorised under ‘university’. The ASAS-COMOSPA study reported 44% university educated participants using the same categorisation. Some indices of country-level socioeconomic status (e.g., HDI and its limited range) may have weaknesses, but results were supported by other indices. Some disease features (e.g., uveitis and peripheral arthritis) were expected to impact work productivity, which was not observed in the current analysis. This may reflect a limitation of the data, which assessed history of each disease feature without indication of severity; for example, an isolated episode of acute anterior uveitis many years ago is unlikely to influence current work productivity. Further, impact of peripheral manifestations may also be captured by ASDAS, BASDAI or BASFI.

In summary, individual- (lower education) and country-level socioeconomic factors (lower healthcare expenditure) were both associated with (lower) work participation. These associations did not differ between SpA phenotypes, but the gender-employment association did vary according to country-level socioeconomic factors (greater male vs female discrepancy in the odds of employment in countries with lower (compared to higher) socioeconomic development). This highlights the need for wider societal interventions, such as improving education and healthcare investment, to improve work outcomes.

## Data Availability

Data from the ASAS-perSpA study are available to investigators on reasonable request. For information on how to access data, contact the Assessment of SpondyloArthritis international Society (www.asas-group.org).

## Funding

None for the current analysis. The ASAS-perSpA study was conducted under the umbrella of ASAS with unrestricted grant of Abbvie, Pfizer, Lilly, Novartis, UCB, Janssen and Merck. The funders did not have any role in the design and conduct of the study; collection, management, analysis and interpretation of the data; preparation, review or approval of the manuscript and decision to submit the manuscript for publication.

## Disclosures

The authors declare no conflicts of interest in relation to this work.

## Contribution

SR, EN and AB designed the study. SSZ analysed the data and all authors were involved in the interpretation and discussion of the results. SSZ wrote the manuscript, with significant input from all co-authors.

## Data availability

Supplementary materials available from reference [16] and the corresponding author. Data from the ASAS-perSpA study are available to investigators on reasonable request. For information on how to access data, contact the Assessment of SpondyloArthritis international Society (www.asas-group.org).

## Acknowledgements

We would like to thank all the collaborators who participated in the study: **José Maldonado-Cocco** (Buenos Aires University School of Medicine, Buenos Aires, Argentina), Hernán Maldonado Ficco (Hospital San Antonio de Padua, Rio Cuarto, Argentina), Rodolfo Pérez Alamino (Hospital Dr. Nicolás Avellaneda, Tucumán, Argentina), Emilio Buschiazzo (Hospital Señor del Milagro, Salta, Argentina), Romina Calvo (Hospital Provincial Dr. José M. Cullen, Santa Fé, Aregntina), Vanesa Duarte (Clínica Monte Grande, Buenos Aires, Argentina), Maria Victoria Martire (Instituto Médico Platense, La Plata, Argentina), Diego Baenas (Hospital Privado de Córdoba, Córdoba, Argentina), Dora Pereira (Hospital Ricardo Gutiérrez, La Plata, Argentina), Adrian Salas (Consultorio Reumatológico, La Plata, Argentina), Juan Manuel Bande (Hospital General de Agudos Dr. E Tornú, Buenos Aires, Argentina), Alberto Berman (Centro Médico Privado de Tucumán, Tucumán, Argentina), **Walter P Maksymowych** (University of Alberta, Canada), Stephanie Belton (University of Alberta, Canada), **Sebastián Ibáñez** (Facultad de Medicina Clínica Alemana – Universdidad del Desarrollo, Santiago de Chile, Chile), María Paz Poblete (Facultad de Medicina Clínica Alemana – Universidad del Desarrollo, Santiago de Chile, Chile), Francisca Valenzuela (Facultad de Medicina Clínica Alemana – Universidad del Desarrollo, Santiago de Chile, Chile), **Wilson Bautista-Molano** (University Hospital Fundación Santa Fé de Bogotá, Bogotá, Colombia), **Jieruo Gu** (Third Affiliated Hospital of Sun Yat-Sen University, Guangzhou, China), Min Xiao (Third Affiliated Hospital of Sun Yat-Sen University, Guangzhou, China), CS Lau (Hong-Kong University, China), Ho Yin Chung (Hong-Kong University, China), **Bassel Elzorkany** (Cairo University, Cairo, Egypt), Sherif Gamal (Cairo University, Cairo, Egypt), Catherine Lebourlout (Cochin Hospital, Paris, France), Daniel Wendling (CHU Besançon, Besançon, France), Clément Prati (CHU Besançon, Besançon, France), Frank Verhoeven (CHU Besançon, Besançon, France), Martin Soubrier (CHU Clermont-Ferrand, Clermont-Ferrand, France), Carine Savel (CHU Clermont-Ferrand, Clermont-Ferrand, France), Trigui Alia (CHU Clermont-Ferrand, Clermont-Ferrand, France), Fan Angélique (CHU Clermont-Ferrand, Clermont-Ferrand, France), Pascal Claudepierre (Henri Mondor Hospital, Créteil, France), Valerie Farrenq (Henri Mondor Hospital, Créteil, France), Kamelia Faramarz (Henri Mondor Hospital, Créteil, France), **Uta Kiltz** (Rheumazentrum Ruhrgebiet, Herne, Germany), Isabella Sieber (Rheumazentrum Ruhrgebiet, Herne, Germany), Dories Morzeck (Rheumazentrum Ruhrgebiet, Herne, Germany), Fabian Proft (Charité University, Berlin, Germany), **Pál Geher** (Semmelweis University, Budapest, Hungary), Edit Toth (Flór Ferenc Hospital, Kistarcsa, Hungary), Katalin Nagy (Markhot Ferenc Hospital, Eger, Hungary), Attila Kovacs (MÁV Hospital, Szolnok, Hungary), **Meghna Gavali** (Nizam’s Institute of Medical Sciences, Hyderabad, India), Liza Rajasekhar (Nizam’s Institute of Medical Sciences, Hyderabad, India), Sapan Pandya (Smt NHL Medical College and Sardar Vallabhbhai Patel Hospital and Vedanta Institute of Medical Sciences, Ahmedabad, India), Bhowmik Meghnathi (Sri Sai Siri Hospital and Prathima Institue of Medical Sciences, Karimnagar, India), **Carlomaurizio Montecucco** (Fondazione IRCCS Policlinico San Matteo, Pavia, Italia), Sara Monti (Fondazione IRCCS Policlinico San Matteo, Pavia, Italia), Alessandro Biglia (Fondazione IRCCS Policlinico San Matteo, Pavia, Italia), **Mitsumasa Kishimoto** (Kyorin University School of Medicine, Tokyo, Japan), Akihiko Asahina (The Jikei University School of Medicine, Japan), Masato Okada (St Luke`s International University and Hospital, Japan), Tadashi Okano (Osaka City University, Japan), Yuko Kaneko (Keio University School of Medicine, Japan), Hideto Kameda (Toho University, Japan), Yoshinori Taniguchi (Kochi University, Japan), Naoto Tamura (Juntendo University School of Medicine, Japan), Shigeyoshi Tsuji (National Hospital Organization Osaka Minami Medical Center, Japan), Hiroaki Dobashi (Kagawa University Faculty of Medicine, Japan), Yoichiro Haji (Daido Hospital, Japan), Akimichi Morita (Nagoya City University, Japan), **Nelly Ziade** (Saint-Joseph University, Beirut, Lebanon), Nelly Salloum (Saint-Joseph University, Beirut, Lebanon), **Rubén Burgos-Vargas** (Hospital General de México Eduardo Liceaga, Mexico City, Mexico), Graciela Meza (CLIDITER), Julio Casasola-Vargas (Hospital General de Mexico, Mexico), César Pacheco-Tena (Hospital General Dr. Salvador Zubirán, Chihuahua, Mexico), Greta Reyes-Cordero (Hospital General Dr. Salvador Zubirán, Chihuahua, Mexico), César Ramos-Remus (Unidad de Investigación de Enfermedades Crónico Degenerativas, Jalisco, Mexico), J Dionisio Castillo (Unidad de Investigación de Enfermedades Crónico Degenerativas, Jalisco, Mexico), Laura González-López (Universidad de Guadalajara, Jalisco, Mexico), Iván Gámez-Nava (Unidad de Investigación Biomédica 02, Hospital de Especialidades, Centro Médico Nacional de Occidente, IMSS Guadalajara, Jalisco, Mexico), **Najia Hajjaj-Hassouni (**International University of Rabat (UIR), Rabat, Morocco), Fadoua Allali (University Mohammed V, CHU Ibn Sina, Rabat, Morocco), Hanan Rkain (University Mohammed V, CHU Ibn Sina, Rabat, Morocco), Lahcen Achemlal (University Mohammed V, CHU Ibn Sina, Rabat, Morocco), Taoufik Harzy (University Sidi Mohammed Benabdellah, CHU Hassan II, Fès, Morocco), **Fernando M Pimentel-Santos** (Universidade NOVA de Lisboa, Lisboa, Portugal), Santiago Rodrigues-Manica (Universidade NOVA de Lisboa, Portugal), Agna Neto (Universidade NOVA de Lisboa, Portugal), Jose Marona (Universidade NOVA de Lisboa, Portugal), Mª Joao Gonçalves (Universidade NOVA de Lisboa, Portugal), Ana Filipa Mourao (Universidade NOVA de Lisboa, Portugal), Rita Pinheiro Torres (Universidade NOVA de Lisboa, Portugal), **Ruxandra Schiotis** (Iuliu Hatieganu University of Medicine, Cluj-Napoca, Romania), Simona Rednic (Iuliu Hatieganu University of Medicine, Cluj-Napoca, Romania), Siao-Pin Simon (Iuliu Hatieganu University of Medicine, Cluj-Napoca, Romania), Laura Muntean (Iuliu Hatieganu University of Medicine, Cluj-Napoca, Romania), Ileana Filipescu (Iuliu Hatieganu University of Medicine, Cluj-Napoca, Romania), Maria Tamas (Iuliu Hatieganu University of Medicine, Cluj-Napoca, Romania), Laura Damian (Iuliu Hatieganu University of Medicine, Cluj-Napoca, Romania), Ioana Felea (Iuliu Hatieganu University of Medicine, Cluj-Napoca, Romania), Dana Fodor (Second Medical Clinic, Emergency Conty Hospital, Cluj-Napoca, Romania), **Tae-Jong Kim** (Chonnam National University Medical School and Hospital, South Korea), Hyun-Yi Kook (Chonnam National University Medical School and Hospital, South Korea), Hyun-Ju Jung (Chonnam National University Medical School and Hospital, South Korea), Tae-Hwan Kim (Hanyang University Hospital for Rheumatic Diseases, South Korea), **Victoria Navarro-Compan** (University Hospital La Paz, Madrid, Spain), Mireia Moreno (Hospital Parc Taulí, Barcelona, Spain), Eduardo Collantes-Estévez (Hospital Universitario Reina Sofía de Córdoba, Spain), M. Carmen Castro-Villegas (Hospital Universitario Reina Sofía, Córdoba, Spain), Cristina Fernández-Carballido (Hospital Universitario San Juan de Alicante, Alicante, Spain), Elizabeth Fernández (Hospital Universtario La Paz, Madrid, Spain), Marta Arévalo (Hospital Parc Taulí, Barcelona, Spain), **Shue-Fen Luo** (Chang Gung Memorial Hospital-Linkou, Taoyuan, Taiwan), Yeong-Jian Jan Wu (Chang Gung Memorial Hospital at Kee-Lung, Taiwan), Tian-Tsai Cheng (Chang Gung Memorial Hospital at Kao-Hsiung, Taiwan), Cheng-Chung Wei (Chung Sun Medical University, Taiwan), **Tuncay Duruöz** (Marmara University School of Medicine, Istanbul, Turkey), Servet Akar (Izmir Katip Çelebi University School of Medicine, Turkey), Ilhan Sezer (Akdeniz University School of Medicine), Umut Kalyoncu (Hacettepe University School of Medicine, Turkey), Sebnem Ataman (Ankara University School of Medicine, Turkey), Meltem Alkan Melikoglu (Erzurum Atatürk University School of Medicine, Turkey), Sami Hizmetli (Sivas Cumhuriyet University School of Medine, Turkey), Ozgur Akgul (Manisa Celal Bayar University School of Medicine, Turkey), Nilay Sahin (Balikesir University School of Medicine, Turkey), Erhan Capkin (Karadeniz Teknik University School of Medicine, Turkey), Fatima Gluçin Ural (Ankara Yildirim Beyazit University School of Medicine, Turkey), Figen Yilmaz (Istanbul Sisli Etfal Training and Research Hospital), Ilknur Aktas (Istanbul Fatih Sultan Mehmet Training and Research Hospital, Turkey), **Floris van Gaalen** (Leiden University Medical Center, The Netherlands), Anne Boel (Leiden University Medical Center, The Netherlands), Mirian Starmans-Kool (Zuyderland Medical Center, The Netherlands), Femke Hoekstra-Drost (Zuyderland Medical Center, The Netherlands), Maha Abdelkadir (Maasstad Hospital in Rotterdam, The Netherlands), Angelique Weel (Maasstad Hospital in Rotterdam, The Netherlands), **Pedro M. Machado** (University College of London, London, UK), **Marina Magrey** (Cases Western Reserve University School of Medicine, Cleveland, Ohio, United States), Darerian Schueller (Cases Western Reserve University School of Medicine, Cleveland, Ohio, United States).

## Steering committee

Joaquim Sieper (Charité University, Berlin, Germany), Desirée van der Heijde (Leiden University Medical Center, The Netherlands), Robert Landewé ((Zuyderland Medical Center, The Netherlands), Anna Moltó (Cochin Hospital, Paris, France).

## Notes

### Competing Interest Statement

The authors have declared no competing interest.

### Author Declarations

Ethical approval was granted by ethical committees in all countries: Argentina, Comite de Docencia e Investigaciones Hospital de Clinicas Dr. Nicolas Avellaneda; Canada, Health Reserach Ethics Board University of Alberta and Alberta Health Services; Chile, Comite etico-Cientifico Servicio Salud Metropolitano Sur Oriente; China, Ethics committee Third Affiliated Hospital of Sun Yat-sen University; Colombia, Comite de Etica en Investigacion del Hospital Militar Central; Egypt, Research Ethics Committee Cairo University Faculty of Medicine; France, Comite de protection des personnes Ile de France III; Germany, Ethics Committee from the medical council Westphalia-Lippe and the Westphalian Wilhelms University; Hungary, Ethics Committee Semmelweis Egyetem Hospital; India, NHL Institutional Review Board (NHLIRB). SMT NHL Municipal Medical College; Italy, Comitato Etico Pavia; Japan, Ethics Committee St Luke's International University; Lebanon, Comite d'ethique Hotel-Dieu de France; Mexico, Comite de Investigacion Hospital General de Mexico Eduardo Liceaga; Morocco, Comite d'Ethique pour la Recherche Biomedicale de Rabat; The Netherlands, Commissie Medische Ethiek Leids Universitair Medisch Centrum; Portugal, Comissao de Etica para a Saude do Centro Hospitalar de Lisboa Ocidental; Romania, Comisia de Etica UMF Iuliu Hatieganu Cluj Napoca; South Korea, Ethics Committee Chonnam National University Medical School from Gwangju; Spain, Comite de Etica de la Investigacion con Medicamentos, Hospital Universitario La Paz; Taiwan, Chang Gung Medical Foundation Institutional Review Board; Turkey, Marmara University School of Medicine Clinical Research Ethics Committee; US, Metrohealth Institutional Review Board. Written informed consent was obtained from all subjects.

