## supplementary for "Association between individual and country-level socioeconomic factors and work participation in spondyloarthritis including psoriatic arthritis: analysis of the ASAS-perSpA study"

| Table S1. Country-level socioeconomic factors for each country. | | | | |
| --- | --- | --- | --- | --- |
| Country | HDI 2018 | HCE 2017 (% GDP) | GDP-PPP 2018 (international$) | Unemployment 2018 (% total labour force) |
| Argentina | 0.8 | 9.1 | 1036 | 9.2 |
| Canada | 0.9 | 10.6 | 1855.8 | 5.8 |
| Chile | 0.8 | 9 | 463.8 | 7.2 |
| China | 0.8 | 5.2 | 21730.7 | 4.3 |
| Colombia | 0.8 | 7.2 | 743 | 9.1 |
| Egypt | 0.7 | 5.3 | 1145.1 | 11.6 |
| France | 0.9 | 11.3 | 3121 | 9.1 |
| Germany | 0.9 | 11.2 | 4514.8 | 3.4 |
| Hungary | 0.8 | 6.9 | 308.7 | 3.7 |
| India | 0.6 | 3.5 | 8995.1 | 5.3 |
| Italy | 0.9 | 8.8 | 2587 | 10.6 |
| Japan | 0.9 | 10.9 | 5230.2 | 2.4 |
| Lebanon | 0.7 | 8.2 | 109.4 | 6.1 |
| Mexico | 0.8 | 5.5 | 2573.8 | 3.3 |
| Morocco | 0.7 | 5.2 | 278.6 | 9.1 |
| Netherlands | 0.9 | 10.1 | 991.9 | 3.8 |
| Portugal | 0.9 | 9 | 353.2 | 7 |
| Romania | 0.8 | 5.2 | 568.9 | 4.2 |
| South Korea | 0.9 | 7.6 | NA | 3.8 |
| Spain | 0.9 | 8.9 | 1894.5 | 15.3 |
| Taiwan | NA | NA | NA | NA |
| Turkey | 0.8 | 4.2 | 2316.4 | 10.9 |
| USA | 0.9 | 17.1 | 20580.2 | 3.9 |
| Human Development Index (HDI) ranks countries according to life expectancy, education and gross national income; Health Care Expenditure (HCE) as percentage of Gross Domestic product (GDP); GDP (based on purchasing power parity); NA not available | | | | |

| Table S2. Participants characteristics (individual socioeconomic factors) for each participating country. | | | | | | | | | |
| --- | --- | --- | --- | --- | --- | --- | --- | --- | --- |
| Country | N | Age, mean (SD) | Male, n (%) | Education | | | Marital status | | |
|  |  |  |  | Primary | Secondary | University | Single | Married | Divorced or Widowed |
| Argentina | 165 | 47.2 (10.6) | 101 (61%) | 23 (14%) | 71 (43%) | 71 (43%) | 39 (24%) | 102 (62%) | 24 (15%) |
| Canada | 11 | 46.6 (9.3) | 9 (82%) | 2 (18%) | 4 (36%) | 5 (45%) | 1 (9%) | 10 (91%) | 0 (0%) |
| Chile | 53 | 45.5 (10.3) | 26 (49%) | 11 (21%) | 28 (53%) | 14 (26%) | 16 (30%) | 27 (51%) | 10 (19%) |
| China | 165 | 31.7 (9.2) | 122 (74%) | 5 (3%) | 75 (45%) | 85 (52%) | 80 (48%) | 82 (50%) | 3 (2%) |
| Colombia | 35 | 45.5 (13.0) | 22 (63%) | 4 (11%) | 11 (31%) | 20 (57%) | 7 (20%) | 23 (66%) | 5 (14%) |
| Egypt | 246 | 40.0 (11.5) | 125 (51%) | 25 (10%) | 83 (34%) | 138 (56%) | 47 (19%) | 176 (72%) | 23 (9%) |
| France | 273 | 42.9 (11.2) | 159 (58%) | 26 (10%) | 113 (42%) | 133 (49%) | 65 (24%) | 183 (67%) | 24 (9%) |
| Germany | 279 | 42.1 (11.6) | 170 (61%) | 6 (2%) | 185 (66%) | 88 (32%) | 82 (29%) | 176 (63%) | 21 (8%) |
| Hungary | 84 | 46.5 (11.8) | 61 (73%) | 14 (17%) | 39 (46%) | 31 (37%) | 24 (29%) | 52 (62%) | 8 (10%) |
| India | 180 | 35.2 (11.0) | 144 (80%) | 21 (12%) | 69 (38%) | 90 (50%) | 71 (39%) | 108 (60%) | 1 (1%) |
| Italy | 150 | 49.5 (10.3) | 71 (47%) | 56 (37%) | 64 (43%) | 30 (20%) | 26 (17%) | 107 (71%) | 17 (11%) |
| Japan | 117 | 47.9 (10.0) | 71 (61%) | 4 (3%) | 63 (54%) | 50 (43%) | 31 (26%) | 71 (61%) | 15 (13%) |
| Lebanon | 177 | 44.1 (11.4) | 104 (59%) | 34 (19%) | 62 (35%) | 81 (46%) | 47 (27%) | 129 (73%) | 1 (1%) |
| Mexico | 189 | 41.8 (12.0) | 121 (64%) | 26 (14%) | 88 (47%) | 74 (39%) | 60 (32%) | 115 (61%) | 14 (7%) |
| Morocco | 251 | 41.2 (11.9) | 157 (63%) | 83 (33%) | 115 (46%) | 53 (21%) | 86 (34%) | 154 (61%) | 11 (4%) |
| Netherlands | 95 | 45.1 (11.8) | 51 (54%) | 1 (1%) | 56 (59%) | 38 (40%) | 10 (11%) | 81 (86%) | 3 (3%) |
| Portugal | 144 | 44.5 (10.3) | 68 (47%) | 36 (25%) | 55 (38%) | 53 (37%) | 36 (25%) | 96 (67%) | 12 (8%) |
| Romania | 116 | 44.9 (11.7) | 80 (69%) | 8 (7%) | 65 (56%) | 43 (37%) | 23 (20%) | 81 (70%) | 12 (10%) |
| South Korea | 195 | 36.9 (11.8) | 151 (77%) | 2 (1%) | 60 (31%) | 133 (68%) | 100 (51%) | 94 (48%) | 1 (1%) |
| Spain | 180 | 45.6 (11.3) | 113 (63%) | 43 (24%) | 66 (37%) | 71 (39%) | 46 (26%) | 117 (65%) | 17 (9%) |
| Taiwan | 191 | 41.7 (11.3) | 139 (73%) | 3 (2%) | 61 (32%) | 127 (66%) | 78 (41%) | 102 (53%) | 11 (6%) |
| Turkey | 455 | 41.2 (10.3) | 242 (53%) | 156 (34%) | 153 (34%) | 146 (32%) | 90 (20%) | 344 (76%) | 21 (5%) |
| USA | 84 | 47.6 (11.1) | 40 (48%) | 1 (1%) | 49 (58%) | 34 (40%) | 33 (39%) | 39 (46%) | 12 (14%) |

| Table S3. Work outcomes for each participating country. | | | | | |
| --- | --- | --- | --- | --- | --- |
| Country |  | Employed | Absenteeism, mean (SD) | Presenteeism, mean (SD) | Overall work impairment, mean (SD) |
|  | N | 3843 | 2253 | 2155 | 2154 |
| Argentina | 165 | 114 (69%) | 6.2 (17.6) | 25.3 (27.5) | 27.2 (28.6) |
| Canada | 11 | 8 (73%) | 0.0 (0.0) | 20.0 (13.1) | 20.0 (13.1) |
| Chile | 53 | 26 (49%) | 21.6 (36.9) | 40.5 (28.7) | 43.3 (30.0) |
| China | 165 | 123 (75%) | 4.3 (12.5) | 18.0 (18.2) | 20.5 (20.2) |
| Colombia | 35 | 25 (71%) | 8.7 (16.4) | 44.8 (23.7) | 47.3 (26.1) |
| Egypt | 246 | 143 (58%) | 22.1 (32.9) | 35.3 (23.1) | 41.4 (27.4) |
| France | 273 | 200 (73%) | 11.3 (28.6) | 27.7 (28.6) | 28.8 (29.7) |
| Germany | 279 | 196 (70%) | 13.3 (30.6) | 28.2 (25.6) | 29.9 (27.2) |
| Hungary | 84 | 62 (74%) | 2.4 (7.9) | 22.5 (25.4) | 23.2 (26.1) |
| India | 180 | 73 (41%) | 17.7 (26.5) | 35.1 (25.7) | 42.8 (30.2) |
| Italy | 150 | 97 (65%) | 7.4 (21.8) | 24.9 (25.7) | 26.4 (27.1) |
| Japan | 117 | 89 (76%) | 1.6 (7.8) | 16.9 (23.5) | 17.8 (24.3) |
| Lebanon | 177 | 121 (68%) | 3.2 (11.3) | 19.9 (25.4) | 21.3 (27.4) |
| Mexico | 189 | 114 (60%) | 8.5 (16.7) | 27.4 (27.4) | 31.0 (30.0) |
| Morocco | 251 | 97 (39%) | 15.2 (32.7) | 30.4 (19.6) | 32.7 (22.1) |
| Netherlands | 95 | 70 (74%) | 15.0 (31.2) | 28.3 (27.1) | 31.1 (30.4) |
| Portugal | 144 | 109 (76%) | 8.0 (21.9) | 27.2 (26.8) | 28.8 (28.0) |
| Romania | 116 | 65 (56%) | 9.8 (25.2) | 23.4 (27.6) | 25.8 (29.6) |
| South Korea | 195 | 103 (53%) | 2.4 (10.7) | 21.0 (22.8) | 21.4 (23.6) |
| Spain | 180 | 129 (72%) | 8.0 (24.8) | 23.0 (30.2) | 23.7 (31.0) |
| Taiwan | 191 | 153 (80%) | 2.8 (10.9) | 28.6 (23.7) | 29.5 (24.5) |
| Turkey | 455 | 219 (48%) | 6.0 (16.0) | 29.2 (26.0) | 31.4 (27.5) |
| USA | 84 | 52 (62%) | 7.5 (24.7) | 24.1 (23.0) | 24.7 (23.4) |

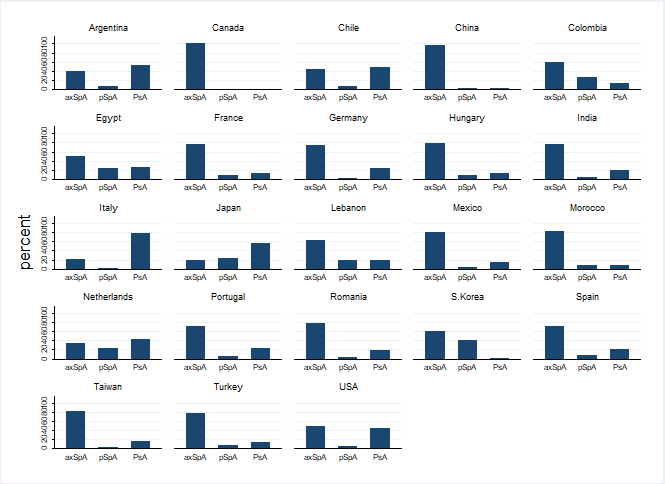

Figure S1. Spondyloarthritis phenotype for each contributing country. axSpA, axial spondyloarthritis; pSpA, peripheral SpA; PsA, psoriatic arthritis.

| Table S4. Participant characteristics according to employment status. | | | | |
| --- | --- | --- | --- | --- |
|  | | Unemployed | Employed | p-value |
| N | | 1447 | 2388 |  |
| Age, years | | 43.4 (13.5) | 41.6 (10.5) | <0.001 |
| Disease duration, years | | 13.7 (11.0) | 13.5 (10.0) | 0.62 |
| Diagnostic delay, years | | 6.1 (8.2) | 5.9 (7.9) | 0.45 |
| Male | | 699 (48%) | 1648 (69%) | <0.001 |
| Education | Primary | 357 (25%) | 233 (10%) | <0.001 |
|  | Secondary | 670 (46%) | 965 (40%) |  |
|  | University | 419 (29%) | 1189 (50%) |  |
| Marital status | Single | 436 (30%) | 662 (28%) | 0.016 |
|  | Married | 893 (62%) | 1576 (66%) |  |
|  | Divorced or Widowed | 116 (8%) | 150 (6%) |  |
| BMI, mean (SD) | | 26.7 (6.1) | 26.2 (5.0) | 0.003 |
| Ever smoking | | 543 (38%) | 1075 (45%) | <0.001 |
| HLA-B27 | Positive | 590 (41%) | 1297 (54%) | <0.001 |
|  | Negative | 336 (23%) | 522 (22%) |  |
|  | Missing | 521 (36%) | 569 (24%) |  |
| SpA phenotype | Axial SpA | 917 (63%) | 1652 (69%) | <0.001 |
|  | Peripheral SpA | 176 (12%) | 224 (9%) |  |
|  | Psoriatic arthritis | 354 (24%) | 512 (21%) |  |
| Axial involvement | | 1127 (78%) | 1918 (80%) | 0.071 |
| Peripheral arthritis | | 859 (59%) | 1222 (51%) | <0.001 |
| Enthesitis | | 696 (48%) | 1012 (42%) | <0.001 |
| Dactylitis | | 195 (13%) | 380 (16%) | 0.040 |
| Uveitis | | 223 (15%) | 419 (18%) | 0.086 |
| Psoriasis | | 401 (28%) | 610 (26%) | 0.14 |
| IBD | | 57 (4%) | 96 (4%) | 0.90 |
| BASDAI | | 4.4 (2.5) | 3.6 (2.4) | <0.001 |
| ASDAS | | 2.9 (1.2) | 2.4 (1.1) | <0.001 |
| CRP, mg/L | | 15.3 (33.6) | 11.1 (37.8) | <0.001 |
| Fatigue (BASDAI Q1) | | 5.1 (2.8) | 4.4 (2.8) | <0.001 |
| BASFI | | 3.8 (2.9) | 2.4 (2.3) | <0.001 |
| Depression/anxiety | | 791 (55%) | 944 (40%) | <0.001 |
| Fibromyalgia diagnosis | | 196 (14%) | 160 (7%) | <0.001 |
| NSAIDs past month | | 1065 (74%) | 1612 (68%) | <0.001 |
| csDMARDs since diagnosis | | 991 (68%) | 1490 (62%) | <0.001 |
| bDMARDs since diagnosis | | 824 (57%) | 1469 (62%) | 0.005 |
| Current prednisolone intake | | 207 (14%) | 198 (8%) | <0.001 |
| All results shown as n (%) or mean (SD), with comparison using chi-squared and t-tests, respectively. Data were incomplete for: education/marital status (n=2), BMI (n=11), BASDAI/Fatigue (n=8), ASDAS (n=47), CRP (n=26), BASFI (n=5), depression (n=8), fibromyalgia (n=2), glucocorticoid use (n=37). IBD, inflammatory bowel disease; BASDAI, Bath AS disease activity index; ASDAS, AS disease activity score; cs/bDMARDs, conventional synthetic/biologic DMARDs. | | | | |

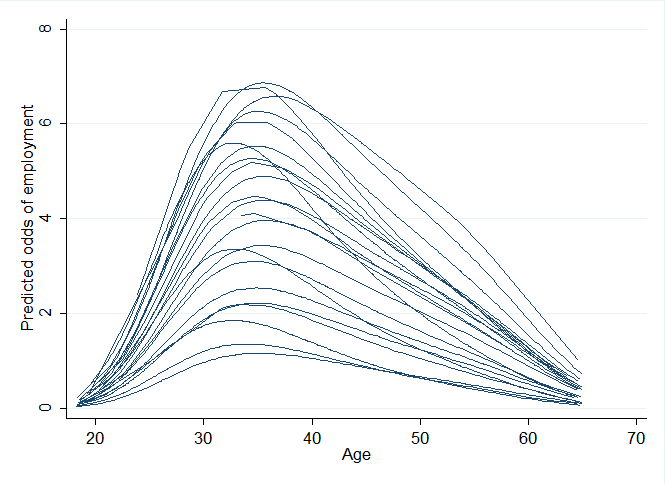

Figure S2. Non-linear (4^th^ degree polynomial) association between age and odds of employment for each country.

| Table S5. Effect of individual socio-economic factors on work outcomes (ASDAS model*) | | | | | |
| --- | --- | --- | --- | --- | --- |
|  | | Employment status  OR (95% CI) | Absenteeism  OR (95% CI) | Presenteeism  OR (95% CI) | Overall work impairment  OR (95% CI) |
| N | | 3780 | 2218 | 2127 | 2126 |
| Age | | 1.43 (1.36,1.51) | 1.00 (0.99,1.01) | 1.00 (0.99,1.01) | 1.01 (1.00,1.02) |
| Age^2^ | | 0.996 (0.995, 0.996) | NS - univariable | NS - univariable | NS - univariable |
| Male (vs female) | | 2.48 (1.92,3.21) | 1.22 (0.96,1.56) | 0.97 (0.78,1.20) | 0.98 (0.80,1.21) |
| Education | Primary | reference | reference | reference | reference |
|  | Secondary | 1.86 (1.48,2.35) | 0.69 (0.49,0.99) | 0.69 (0.49,0.99) | 0.81 (0.58,1.14) |
|  | University | 3.68 (2.87,4.72) | 0.67 (0.47,0.96) | 0.49 (0.34,0.69) | 0.57 (0.41,0.81) |
| Marital status | Single | reference | reference | reference | reference |
|  | Married | 1.27 (1.04,1.56) | 0.95 (0.73,1.22) | 0.98 (0.78,1.22) | 0.99 (0.80,1.23) |
|  | Divorced or Widowed | 1.39 (0.98,1.97) | 1.39 (0.88,2.18) | 1.16 (0.74,1.82) | 1.26 (0.81,1.95) |
| ASDAS | | 0.78 (0.72,0.84) | 1.51 (1.33,1.72) | 2.31 (2.04,2.61) | 2.02 (1.79,2.27) |
| Fatigue (BASDAI Q1) | | NS - multivariable | 1.14 (1.09,1.21) | 1.30 (1.24,1.36) | 1.30 (1.24,1.36) |
| Depression/anxiety | | 0.70 (0.59,0.82) | 1.45 (1.15,1.82) | 1.95 (1.59,2.39) | 1.86 (1.52,2.27) |
| Fibromyalgia | | NS - multivariable | 1.62 (1.11,2.35) | 1.58 (1.03,2.41) | 1.91 (1.26,2.89) |
| BMI | | NS - multivariable | 0.99 (0.97,1.02) | NS - multivariable | NS - multivariable |
| Dactylitis | | 1.41 (1.12,1.76) | NS - univariable | NS - univariable | NS - univariable |
| Uveitis | | NS - multivariable | 0.62 (0.46,0.84) | NS - univariable | NS - univariable |
| NSAIDs | | NS - multivariable | 1.53 (1.18,1.99) | 1.32 (1.07,1.64) | 1.26 (1.03,1.55) |
| bDMARDs | | NS - multivariable | NS - univariable | 1.23 (1.01,1.51) | NS - univariable |
| Results from multivariable mixed effects logistic (employment) or ordinal logistic (absenteeism, presenteeism, overall work impairment) models. *Models used ASDAS/fatigue rather than BASDAI or BASFI, which were collinear thus modelled separately.  ASDAS, AS disease activity score; bDMARD, biologic DMARD; BMI, body mass index; OR, odds ratio; 95%CI, 95% confidence interval; NS, not significant (at P<0.05) at the univariable or multivariable modelling stage. | | | | | |

| Table S6. Effect of individual socio-economic factors on work outcomes (BASFI model*). | | | | | |
| --- | --- | --- | --- | --- | --- |
|  | | Employment status  OR (95% CI) | Absenteeism  OR (95% CI) | Presenteeism  OR (95% CI) | Overall work impairment  OR (95% CI) |
| N | | 3820 | 2225 | 2149 | 2127 |
| Age | | 1.46 (1.38,1.53) | 0.99 (0.98,1.00) | 0.99 (0.98,1.00) | 0.99 (0.98,1.00) |
| Age^2^ | | 0.996 (0.995, 0.996) | NS - univariable | NS - univariable | NS - univariable |
| Male (vs female) | | 2.61 (2.00,3.40) | 1.15 (0.90,1.47) | 0.98 (0.79,1.21) | 0.95 (0.77,1.18) |
| Education | Primary | reference | reference | reference | reference |
|  | Secondary | 1.87 (1.47,2.36) | 0.68 (0.47,0.97) | 0.68 (0.47,0.97) | 0.80 (0.56,1.14) |
|  | University | 3.66 (2.85,4.70) | 0.64 (0.44,0.91) | 0.47 (0.33,0.67) | 0.57 (0.40,0.82) |
| Marital status | Single | reference | reference | reference | reference |
|  | Married | 1.25 (1.02,1.54) | 0.94 (0.73,1.22) | 0.94 (0.75,1.17) | 0.96 (0.77,1.20) |
|  | Divorced or Widowed | 1.39 (0.98,1.97) | 1.27 (0.80,2.01) | 1.00 (0.63,1.58) | 1.08 (0.69,1.70) |
| BASFI | | 0.80 (0.77,0.83) | 1.33 (1.27,1.40) | 1.74 (1.63,1.85) | 1.68 (1.58,1.78) |
| Fatigue (BASDAI Q1) | | 1.08 (1.04,1.12) | NS - multivariable | 1.29 (1.23,1.35) | 1.26 (1.20,1.31) |
| Depression/anxiety | | 0.72 (0.61,0.86) | 1.63 (1.31,2.04) | 1.95 (1.59,2.41) | 1.86 (1.52,2.28) |
| Fibromyalgia | | NS - multivariable | 1.62 (1.11,2.36) | NS - multivariable | NS - multivariable |
| Dactylitis | | 1.36 (1.08,1.70) | NS - univariable | NS - univariable | NS - univariable |
| Uveitis | | NS - multivariable | 0.64 (0.48,0.86) | NS - univariable | NS - univariable |
| NSAIDs | | NS - multivariable | 1.73 (1.33,2.26) | 1.28 (1.04,1.59) | 1.27 (1.03,1.56) |
| bDMARDs | | NS - multivariable | 1.33 (1.05,1.67) | NS - multivariable | NS - univariable |
| Glucocorticoids | | NS - multivariable | 1.52 (1.08,2.15) | NS - multivariable | 1.52 (1.07,2.17) |
| Results from multivariable mixed effects logistic (employment) or ordinal logistic (absenteeism, presenteeism, overall work impairment) models. *Models used BASFI/fatigue rather than ASDAS or BASDAI, which were collinear thus modelled separately.  BASFI, Bath AS Functional Index; bDMARD, biologic DMARD; BMI, body mass index; OR, odds ratio; 95%CI, 95% confidence interval; NS, not significant (at P<0.05) at the univariable or multivariable modelling stage. | | | | | |

| Table S7. Effect of individual socio-economic factors on work outcomes (BASDAI model*). | | | | | |
| --- | --- | --- | --- | --- | --- |
|  | | Employment status  OR (95% CI) | Absenteeism  OR (95% CI) | Presenteeism  OR (95% CI) | Overall work impairment  OR (95% CI) |
| N | | 3817 | 2248 | 2151 | 2150 |
| Age | | 1.43 (1.36,1.51) | 0.99 (0.98,1.01) | 1 (0.99,1.01) | 1 (0.99,1.01) |
| Age^2^ | | 0.996 (0.995, 0.996) | NS - univariable | NS - univariable | NS - univariable |
| Male (vs female) | | 2.42 (1.86,3.15) | 1.25 (0.98,1.60) | 0.99 (0.80,1.22) | 1.02 (0.83,1.26) |
| Education | Primary | reference | reference | reference | reference |
|  | Secondary | 1.86 (1.48,2.35) | 0.69 (0.48,0.98) | 0.69 (0.48,0.98) | 0.81 (0.57,1.14) |
|  | University | 3.85 (3.01,4.93) | 0.67 (0.47,0.96) | 0.50 (0.35,0.71) | 0.60 (0.42,0.84) |
| Marital status | Single | reference | reference | reference | reference |
|  | Married | 1.29 (1.05,1.58) | 0.91 (0.71,1.18) | 0.88 (0.71,1.10) | 0.89 (0.72,1.10) |
|  | Divorced or Widowed | 1.37 (0.97,1.94) | 1.42 (0.91,2.24) | 1.17 (0.75,1.82) | 1.21 (0.78,1.87) |
| BASDAI | | 0.92 (0.89,0.96) | 1.37 (1.30,1.44) | 1.96 (1.86,2.08) | 1.85 (1.75,1.95) |
| Depression/anxiety | | 0.67 (0.56,0.79) | 1.44 (1.15,1.81) | 1.91 (1.55,2.34) | 1.80 (1.47,2.20) |
| Fibromyalgia | | NS - multivariable | 1.56 (1.08,2.27) | NS - multivariable | 1.74 (1.14,2.65) |
| Dactylitis | | 1.43 (1.14,1.79) | NS - univariable | NS - univariable | NS - univariable |
| Uveitis | | NS - multivariable | 0.65 (0.49,0.88) | NS - univariable | NS - univariable |
| NSAIDs | | NS - multivariable | 1.57 (1.21,2.05) | 1.33 (1.07,1.65) | 1.23 (1.00,1.52) |
| bDMARDs | | NS - multivariable | NS - multivariable | 1.27 (1.04,1.56) | NS - univariable |
| Results from multivariable mixed effects logistic (employment) or ordinal logistic (absenteeism, presenteeism, overall work impairment) models. *Models used BASDAI/CRP rather than BASFI or BASDAI, which are collinear thus modelled separately.  BASDAI, Bath AS Disease Activity Index; bDMARD, biologic DMARD; BMI, body mass index; OR, odds ratio; 95%CI, 95% confidence interval; NS, not significant (at P<0.05) at the univariable or multivariable modelling stage. | | | | | |
